# Constructing germline research cohorts from the discarded reads of clinical tumor sequences

**DOI:** 10.1101/2021.04.09.21255197

**Authors:** Alexander Gusev, Stefan Groha, Kodi Taraszka, Yevgeniy R. Semenov, Noah Zaitlen

## Abstract

**Background:** Hundreds of thousands of cancer patients have had targeted (panel) tumor sequencing to identify clinically meaningful mutations. In addition to improving patient outcomes, this activity has led to significant discoveries in basic and translational domains. However, the targeted nature of clinical tumor sequencing has a limited scope, especially for germline genetics. In this work, we assess the utility of discarded, off-target reads from tumor-only panel sequencing for recovery of genome-wide germline genotypes through imputation.

**Methods:** We develop a framework for inference of germline variants from tumor panel sequencing, including imputation, quality control, inference of genetic ancestry, germline polygenic risk scores, and HLA alleles. We benchmark our framework on 833 individuals with tumor sequencing and matched germline SNP array data. We then apply our approach to a prospectively collected panel sequencing cohort of 25,889 tumors.

**Results:** We demonstrate high to moderate accuracy of each inferred feature relative to direct germline SNP array genotyping: individual common variants were imputed with a mean accuracy (correlation) of 0.86; genetic ancestry was inferred with a correlation of >0.98; polygenic risk scores were inferred with a correlation of >0.90; and individual HLA alleles were inferred with correlation of >0.89. We demonstrate a minimal influence on accuracy of somatic copy number alterations and other tumor features. We showcase the feasibility and utility of our framework by analyzing 25,889 tumors and identifying relationships between genetic ancestry, polygenic risk, and tumor characteristics that could not be studied with conventional data.

**Conclusions:** We conclude that targeted tumor sequencing can be leveraged to build rich germline research cohorts from existing data, and make our analysis pipeline publicly available to facilitate this effort.

## BACKGROUND

Large-scale targeted tumor sequencing is ubiquitous in the clinical setting, and hundreds of thousands of cancer patients have had tumors sequenced at targeted cancer-related genes in order to identify clinically actionable mutations[1–5]. Data from these cohorts provide an unprecedented opportunity for basic research and translational discovery, improving our understanding of cancer biology and supporting clinical decision-making. Recent FDA approval of such technologies across many cancer types will likely lead to even broader adoption in the coming years[6].

Research involving these cohorts has identified novel somatic mutations that alter risk[7], treatment responses, and outcomes[2,8–11]. However, clinical tumor sequencing platforms focus on exons within a small number of known cancer genes[1,2], thereby excluding potentially actionable germline and/or non-coding variants. In contrast to targeted sequencing, the information obtained from whole-genome sequencing (WGS) provides more comprehensive insights into cancer genetics[12–15], but sequencing costs have limited the sample size and power of these studies.

Recent work has shown that ultra low-coverage sequencing, including off-target regions from targeted sequencing, can be used to accurately impute common germline polymorphisms by leveraging linkage disequilibrium (LD) information within the low-coverage data[16–22]. Here, we demonstrate that similar techniques can be used to infer common germline variation from targeted sequencing of tumors. We first use extensive benchmarking with real tumor/germline data to show that off-target tumor sequencing can be used to accurately estimate common germline genotypes. We then aggregate these genotypes to infer genetic ancestry, polygenic risk scores (PRS), and HLA alleles, and demonstrate high to moderate imputation accuracy of each. Finally, we showcase the research utility of this data by identifying associations with germline risk and genetic ancestry in a “real world” cohort of >25,000 tumors.

## METHODS

### Overview of data

We collected 25,889 tumors spanning >20 cancer types as part of the Dana-Farber PROFILE cohort, which were prospectively sequenced on the OncoPanel platform as part of routine cancer care[23] (**Figure S1a**). The OncoPanel platform targeted the exons of 275-447 cancer genes on one of three panel versions[1,23]. Genomewide, the mean and median sequencing coverage was 0.17x and 0.13x respectively (**Figure S1b**), compared to a mean on-target coverage of ∼200x. A subset of 833 individuals had DNA available from whole blood and were genotyped on the Illumina Multi-Ethnic Genotyping Array (MEGA) and used for benchmarking. Written informed consent was obtained from participants prior to inclusion in this study.

### Patient consent, accrual, and genotyping

PROFILE samples were selected and sequenced from patients who were consented under institutional review board (IRB) approved protocol 11-104 and 17-000 from the Dana-Farber/Partners Cancer Care Office for the Protection of Research Subjects. Written informed consent was obtained from participants prior to inclusion in this study. Secondary analyses of previously collected data were performed with approval from the Dana-Farber IRB (DFCI IRB protocol 19-033 and 19-025; waiver of HIPAA authorization approved for both protocols). Patients were recruited based on available material and consent, and were not otherwise ascertained for age, sex, stage, or tumor site. Each sample was sequenced on one of three panel versions targeting the exons of 275, 300, and 447 genes respectively[1,23]. Sequencing was performed using an Illumina HiSeq 2500 with 2×100 paired-end reads. Samples met a minimum of 30X coverage for 80% of targets for analysis.

A subset of 833 patients were germline genotyped as part of the Mass General Brigham Biobank. DNA samples were processed from whole blood and genotyped on either the Illumina Multi-Ethnic Genotyping Array (MEGA), the Expanded Multi-Ethnic Genotyping Array (MEGA Ex) array, or the Multi-Ethnic Global (MEG) BeadChip. All germline samples were imputed to the Haplotype Reference Consortium reference panel and then restricted to ∼1.1 million HapMap3 variants that typically exhibit high imputation accuracy across genotyping platforms.

### Germline imputation

We assessed two imputation algorithms intended for low-coverage data: STITCH[20] and GLIMPSE[45]. For all analyses, OncoPanel data was aligned to hg19 using bwa and processed with the GATK IndelRealigner. The 1000 Genomes Phase 3 release was used as a haplotype reference, targeting variants with >1% frequency in the European population. For STITCH, aligned reads were passed to the imputation algorithm in 5MB batches; detailed parameters are described in the associated code (see **Data Availability**). For GLIMPSE, the multi-step imputation workflow was applied with default parameters. We considered two other imputation approaches: GeneImp[46] and BEAGLE[47] but found that their computational requirements were infeasible for n>1000. Variants were considered “QC passing” if they had a minor allele frequency >1% and an INFO score >0.4 (similar to parameters used in ref.[19]). Additional QC thresholds were investigated in **Figure S4**.

### Germline HLA allele inference

HLA alleles were inferred from the imputed germline genotypes using the SNP2HLA pipeline and the NIDDK HLA reference panel with default parameters. The NIDDK reference panel[48] contained 5,225 samples with serotyped HLA alleles spanning HLA A, B, C, DPA1, DPB1, DQA1, DQB1, and DRB1. Restricting to alleles with at least 1% carriers, the panel contained 83 2-digit alleles and 112 4-digit alleles. We did not find that additional SNP exclusion or quality control prior to HLA inference produced any measurable improvement in accuracy and thus used all imputed variants retained after the first post-imputation QC step. After HLA imputation, we retained HLA variants with an imputation confidence >0.75 and at least one call with probability >0.5.

HLA imputation yielded an estimate of carrier and non-carrier status for each allele, which does not map directly to homozygosity due to the presence of multiple alleles per locus. HLA homozygosity (*h*) was computed for each individual and locus (A, B, C, and D**\***) as follows:

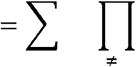

where is the probability of being a homozygous carrier for allele *a* and is the probability of being a non-carrier of allele *b*. The probability of being homozygous for at least one locus was then computed across loci as follows:

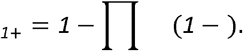

with separate computations of _*1*+_ for the MHC class I and class II alleles.

### Quantifying somatic copy number alterations

Local copy number was called by the default analysis pipeline used for clinical reporting to patients and physicians. The RobustCNV (v2.0.1) algorithm was applied to individual tumors along with a panel of normals to identify copy number segments based on coverage[49]. We then identified the 5% of individual-segment pairs with the highest and lowest estimated segment mean in the population and computed imputation accuracy within these individual-segment pairs or for all other (“neutral”) regions.

### Polygenic risk score inference

Publicly available GWAS data from studies of breast cancer, prostate cancer, ovarian cancer, height, BMI, and smoking status were used to evaluate polygenic risk score and association statistic accuracy [50–52]. Risk scores were restricted to HapMap3 SNPs, which are typically well imputed and capture all of the GWAS SNP-heritability. A risk score for each trait and individual was constructed as the sum of allele dosages weighted by the GWAS association statistic using either (a) off-target imputed SNPs or (b) genotype array SNPs as the gold standard. To limit parameter tuning, no variant pruning nor thresholding was employed and all ∼1.1M GWAS SNPs were used. Accuracy was quantified using the slope and correlation from a linear regression of the off-target score on the gold-standard score.

### Genetic ancestry inference

Samples were projected into genetic ancestry principal components using the weights previously derived by the SNPWEIGHTS software[35] for the continental populations. Weights were constructed from the 1000 Genomes reference groups with ancestry from Northern/Western Europe (CEU), Western Africa (YRI), and China (CHB+CHD). In our data, each component was projected independently as a linear combination of the weights and individual sample dosages (using the plink2 “--score” command). Components were then linearly recalibrated by fitting to self-reported race as an outcome (note this linear recalibration is for interpretation purposes only and does not influence the significance of any downstream associations). To estimate ancestry fractions, we uniformly rescaled the African and Asian components to be between 0-1 and additionally uniformly scaled the ancestry of each individual to be between 0-1.

### Analysis of *EGFR* mutation carriers in NSCLC

We restricted to 2,900 NSCLC samples from the full cohort and quantified carrier status for somatic SNVs in the *EGFR* gene. All samples targeted *EGFR* for sequencing. Somatic variants were called using the default analysis pipeline used for clinical reporting: the MuTect algorithm[53] with a panel of normals followed by filtering of any common variants in the Gnomad reference panel[54]. Only non-synonymous variants in *EGFR* exons were retained and being a carrier was defined as having >0 mutations. Carrier status was associated with genetic ancestry using logistic regression with covariates for sex, age, tumor purity, and panel versions.

## RESULTS

### Accurate inference of common germline genotypes from tumor-only sequencing

Common germline genotypes were imputed directly from off-target tumor sequencing reads (**Figure 1**; see Methods) using the 1000 Genomes reference panel, and evaluated against the gold standard germline SNP genotyping. For comparison across platforms, the SNP array data were also imputed using standard pipelines via the Haplotype Reference Consortium, and both cohorts restricted to ∼1.1 million common SNPs from the HapMap3 panel, which are typically well imputed and reflect common genetic variation. Both cohorts imputed the allele “dosage” for each individual and site, defined as the expected number of non-reference alleles carried by the individual. For each site, accuracy was quantified using: Pearson correlation across individuals; mean error (i.e. the mean difference between the genotyped and imputed variant); and mean absolute error. We evaluated multiple imputation approaches and found that the STITCH algorithm yielded the highest accuracy while able to scale to tens of thousands of samples (see Methods; **Figure S2**).

**Figure 1:**
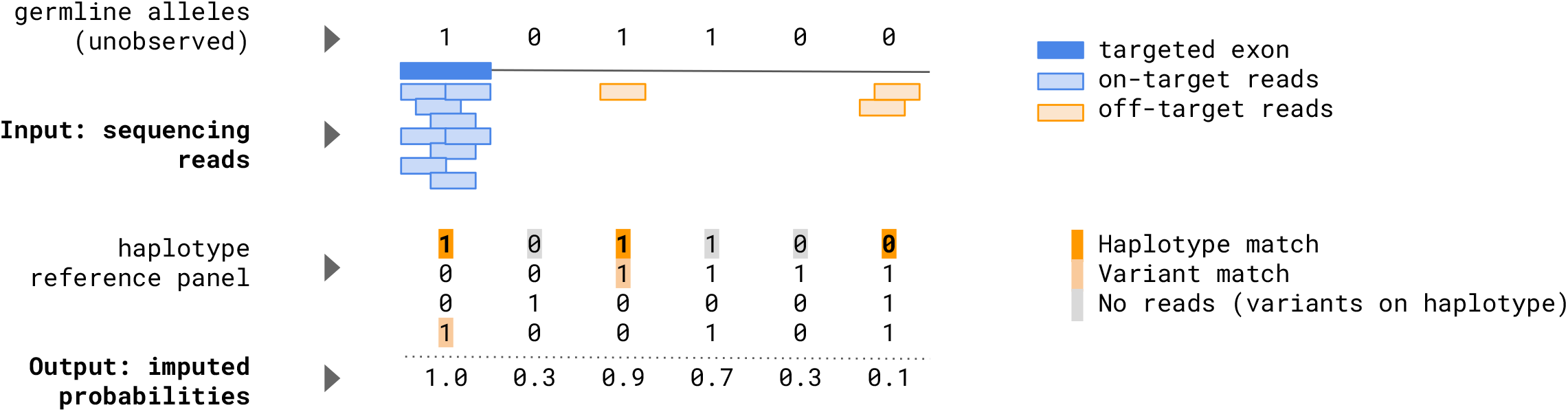
Schematic of germline imputation from low-coverage sequencing. The sequenced genome is shown as a horizontal line with targeted regions (dark blue), on-target reads aligning to these regions (light blue), and off-target reads typically discarded (orange). The haplotype reference panel is shown as a matrix of 0/1 alleles with alleles that match sequenced reads shown in light orange; alleles that form a haplotype match shown in dark orange; and alleles that reside along the haplotype but did not carry reads in the target sample shown in gray. Conceptually, the matched haplotype is used to refine the dark orange alleles and impute the light gray alleles in the target individual. A schematic of the imputed variant probabilities is shown at the bottom. For simplicity, we assume all individuals are haploid and alleles are on the 0-1 scale. In practice, diploid imputation is performed and the resulting alleles are on the 0-2 scale.

Tumor imputed variants exhibited high to moderate correlation with the true germline variant across the entire genome (**Figure 2**). Mean correlation was 0.79 (s.e. 0.0002) across all 1.1 million SNPs and increased to 0.86 (s.e. 0.0001) when restricting to 927,436 variants that passed conventional QC thresholds (see Methods) (**Figure 2a**). A total of 37% of QC passing SNPs had a correlation >0.9 and <0.5% exhibited a correlation of <0.6 (compared to 13% of all SNPs exhibiting a correlation of <0.6) (**Figure 2b**; **Figure S3**). QC filtering, which did not rely on knowledge of the germline genotypes, thus removed primarily low accuracy SNPs, and we restricted to filtered variants for the remainder of our analyses. Imputation confidence (INFO score) was the primary determinant of imputation accuracy and filtering on other parameters did not substantially impact accuracy (**Figure S4**).

**Figure 2:**
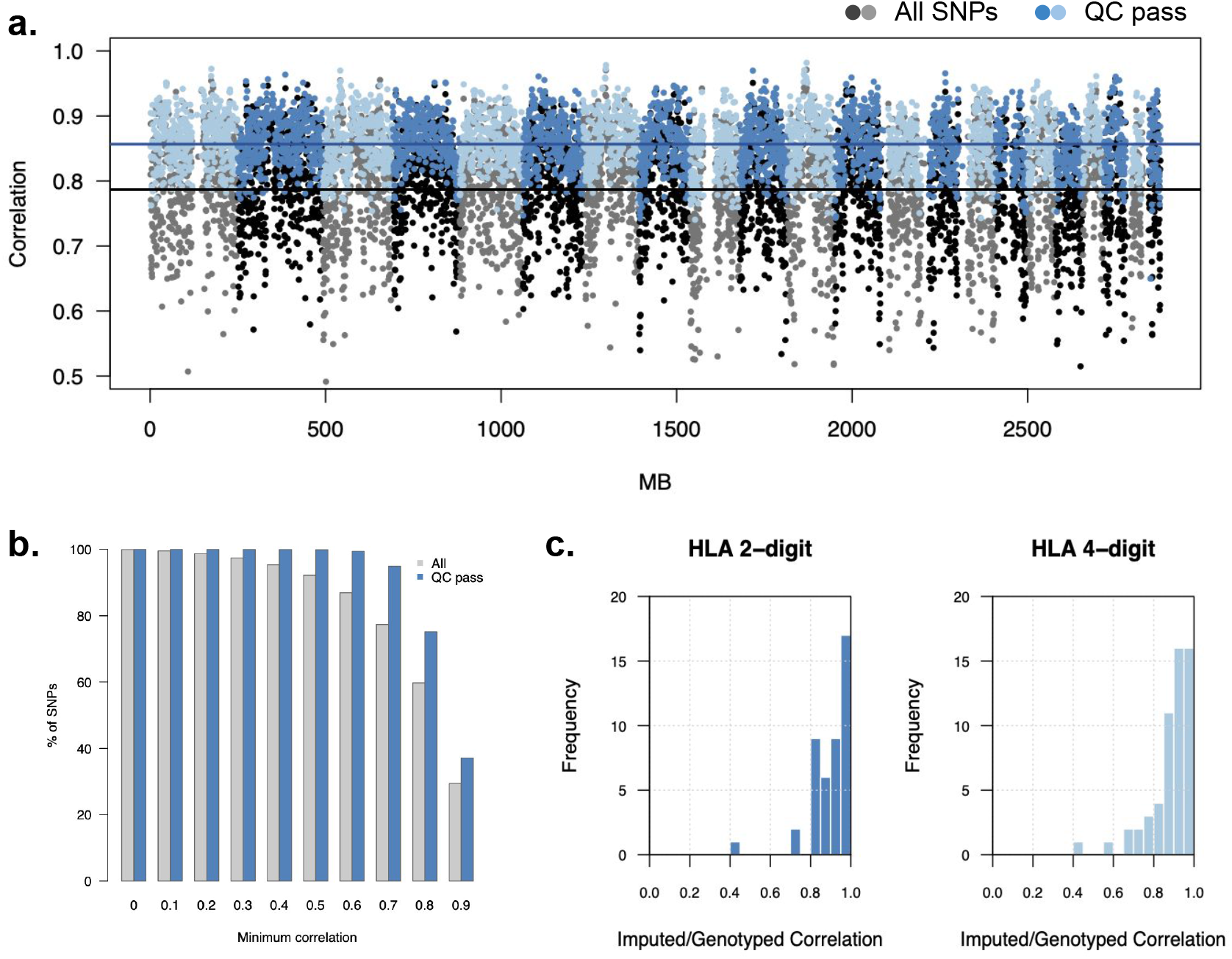
Germline imputation accuracy from tumor sequences. (**a**) Mean imputation accuracy (Pearson correlation; y-axis) across sliding windows of 200 SNPs shown for all variants (gray/black) and QC-passing variants (blue/light blue) with alternating shading for chromosomes. (**b**) Fraction of variants (y-axis) as a function of minimum imputation accuracy (Pearson correlation; x-axis) for all variants (gray) and QC-passing variants (blue). (**c**) Histogram of imputation accuracy (Pearson correlation) for 2-digit and 4-digit HLA alleles after QC.

### Minimal influence of somatic copy number alterations on imputation accuracy

Tumor genomes often harbor extensive somatic alterations that have the potential to influence imputation. We investigated the relationship between somatic copy number alterations (SCNAs) and imputation accuracy by quantifying variant accuracy for the 5% most strongly deleted and the 5% most strongly amplified segments in this cohort (see Methods). As correlation can be highly uncertain (or incalculable) when computed over a small number of individuals, we focused on the allelic error metrics for this analysis. Allelic error was always computed relative to the major allele to capture any systematic directional biases in the imputation. Surprisingly, for the 5% most amplified regions, absolute error decreased relative to the rest of the genome (more sites imputed with zero error) with no visible artifacts (**Figure 3**). This was consistent with amplified regions having higher coverage and thus more reads for the imputation scaffolding, without degrading accuracy. For the most deleted regions, error increased (fewer sites imputed with zero error) and imputed variants exhibited a small but statistically significant bias towards the major allele (**Figure 3**). This again was consistent with deleted regions exhibiting lower coverage and fewer reads for imputation. In sum, extreme SCNAs had a small influence on imputation error, with deletions leading to lower accuracy and a slight bias towards the major allele.

**Figure 3:**
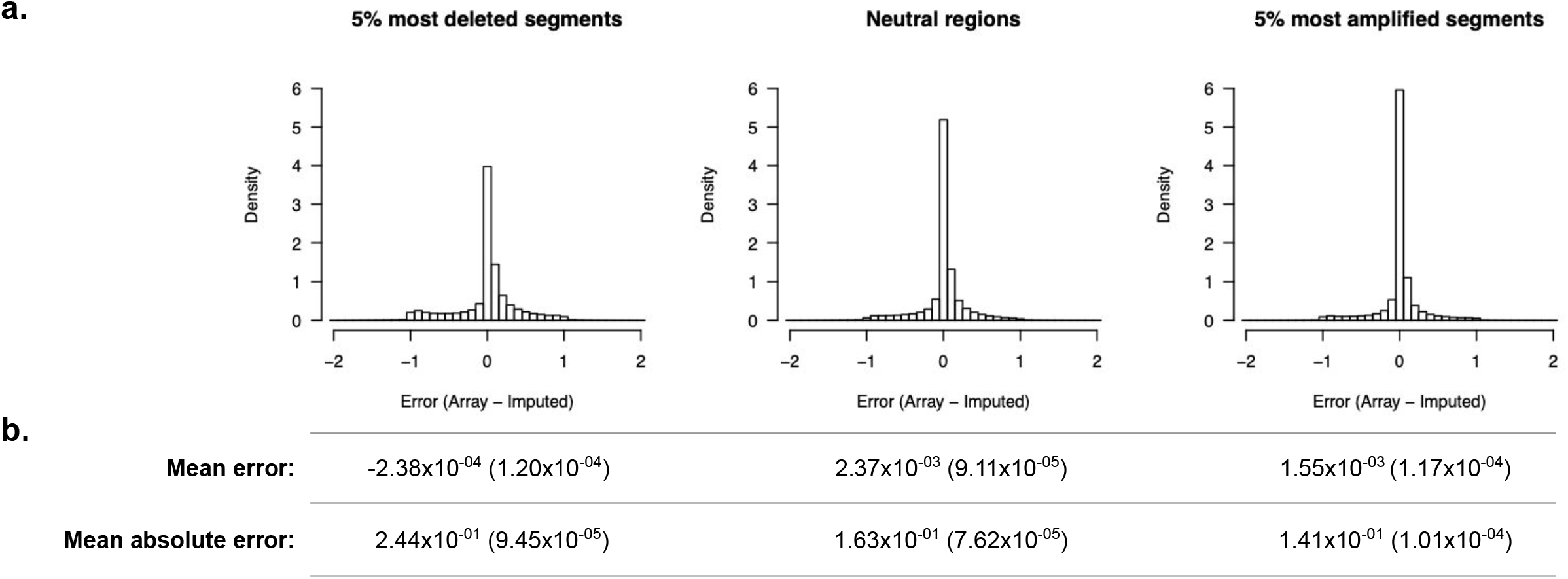
Imputation accuracy for somatically deleted or amplified regions. (**a**) Histogram of imputation error (difference between imputed dosage and true allele) for the individual regions most deleted (left), amplified (right), and the rest (middle). (**b**) Table of mean error and mean absolute error for each region type. Standard error reported in parentheses.

We separately considered the association of tumor-level features with genome-wide imputation error, without restricting to specific regions. The associations were quantified in a multivariate linear regression with mean absolute imputation error as the dependent variable (**Table S1**). Sequencing panel version was by far the most significantly associated feature, explaining 40.6% of the variance in imputation error (p<2×10^−16^ for all panel versions). Beyond panel version, tumor purity was associated with a slight increase in error (increase of 0.006 in error for tumors with >50% purity relative to others, p=0.02; with comparable significance for tumor purity as a quantitative feature) and metastatic tumors were nominally associated with increased error (increase of 0.005 in error, p=0.08). While significant, the effect sizes of these features were minor relative to a mean imputation error of 0.18 (s.d. 0.04) and only increased the overall variance explained from 40.6% to 41.1%. Neither tumor mutational burden nor total copy number burden were significantly associated with error (**Table S1**). We note that somatic single nucleotide variants (SNVs) were unlikely to introduce a bias in imputation because their positions are either absent from the imputation reference panels or present at very low frequency such that no attempt to impute them is made.

### Imputation of germline HLA alleles

Genetic variation at the HLA locus has been associated with multiple cancer-related phenotypes[14,24–26], and we investigated the ability of tumor imputed variants to recover HLA alleles. Importantly, HLA genes were not targeted directly by any of the panels so all inference was based on off-target polymorphisms. A conventional HLA imputation algorithm was used to infer the germline HLA alleles from a reference panel of eight common class I and class II genes[27] using the tumor imputed variants as input (see Methods). As before, we benchmarked against an independent imputation performed from the germline SNP array data. After restricting to high confidence imputed alleles (see Methods), the mean imputed/genotyped correlation was 0.89 (s.e. 0.11) and 0.90 (s.e. 0.10) for four-digit and two-digit resolution, respectively. All but two individual alleles exhibited correlation >0.6: HLA-DRB1**\***10:01 (corr = 0.44) and HLA-C**\***07:04 (corr = 0.57) (**Figure 2c**). Lastly, we used the tumor-imputed alleles to estimate whether an individual is homozygous for at least one HLA allele (_1+_; see Methods), as HLA homozygosity has been hypothesized to be a biomarker of response in some cancers[24,28]. The AUC for _1+_ was 0.98 for MHC class I alleles, and 0.81 for MHC class II alleles (**Figure S5**). Tumor imputed variants can thus be directly used for accurate downstream imputation of HLA alleles.

### Inference of germline polygenic risk scores

Common germline variants are increasingly being used to predict disease risk by aggregating individual effect-sizes into polygenic risk scores (PRSs)[29,30] and we investigated the accuracy of PRSs computed from the tumor-imputed variants. We selected the PRS from a recent large-scale breast cancer GWAS[31] as representative (findings were similar for other PRSs from polygenic traits: **Figure S6**). For each individual, a risk score was computed using the tumor imputed variants and compared to that computed using the germline genotypes. Pearson correlation across individuals for the two scores was 0.92 with no observable outliers, and a slight linear deflation of the score as would be expected from noise due to imputation (**Figure 4a**). We confirmed that PRS imputation error (computed as the difference between the imputed and true PRS) was approximately normally distributed (**Figure 4b**) and consistent across the true PRS deciles (Figure 4c). Lastly, we found no statistically significant difference in the error distribution across cancer types (in a multivariate linear regression; **Figure 4d**), nor was any cancer type individually associated with mean error or mean absolute error. Likewise, no significant mean differences were observed across the three sequencing panel versions (**Figure S7**), nor was there a significant association between tumor purity and error or absolute error. Similar PRS accuracy and results were observed for other common traits (**Figure S6**).

**Figure 4:**
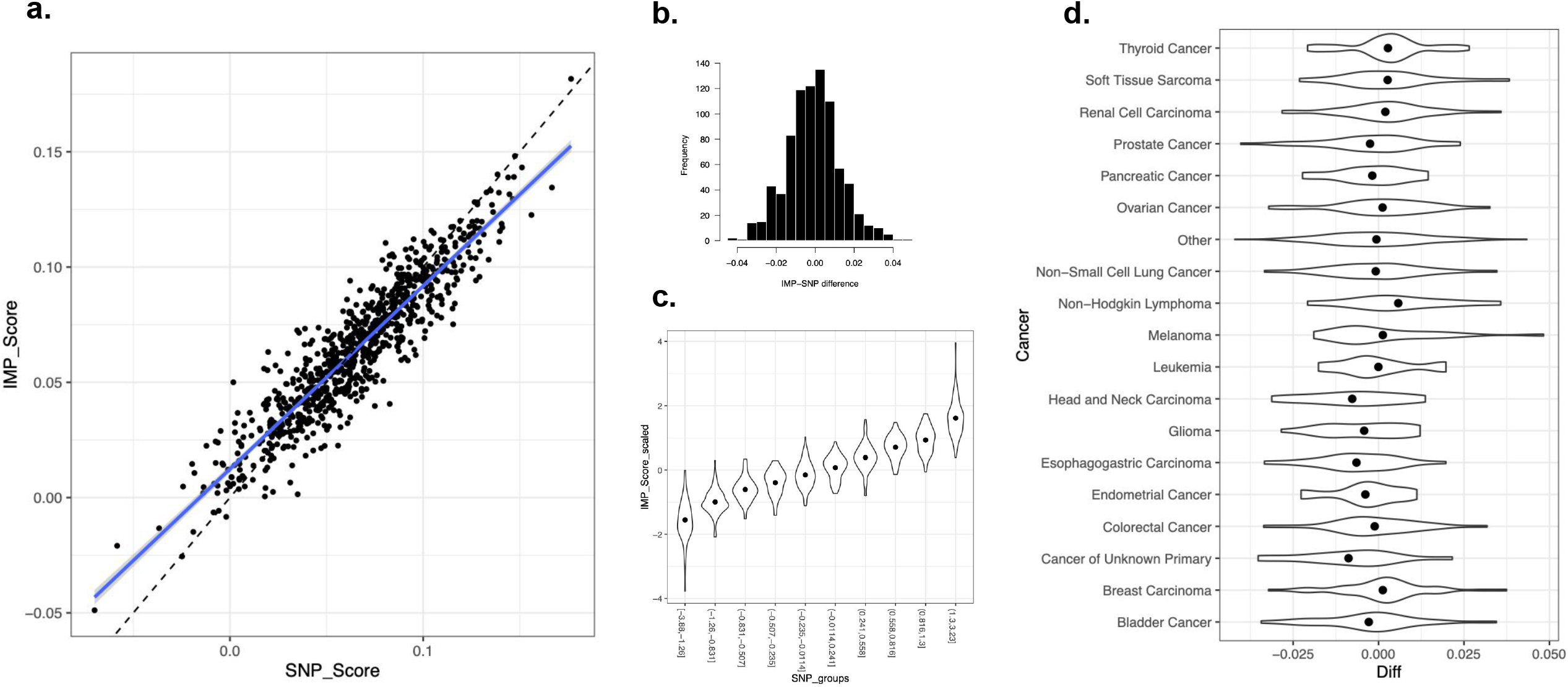
Polygenic Risk Score (PRS) accuracy. (**a**) Scatter plot of germline SNP (x-axis) and tumor imputed (y-axis) polygenic risk score across individuals. Each dot is an individual; dashed line shows y=x diagonal; blue line shows the linear fit. (**b**) Histogram of imputed/genotyped difference across individuals. (**c**) Violin plot of imputed score density (y-axis) as a function of genotyped score decile (x-axis). (**d**) Violin plot of the density of imputed/genotyped score differences (x-axis) across cancer types (y-axis).

Having established the high accuracy of the tumor imputed PRS we applied it to the full cohort of tumors to investigate differences between cancers. We constructed PRSs for common cancers (breast, glioma, lung, and melanoma) as well as risk PRSs for exposures typically associated with these cancers (smoking and tanning), using a simple pruning and thresholding approach[32] (see Methods). We note that a PRS for an exposure is a prediction of the genetic predisposition for the corresponding behavior (e.g. propensity to smoke) and not a direct measurement of the exposure. We then tested each score for association with cancer type, with cases defined as patients having a focal cancer and controls defined as patients with any other cancer (note, no genuine healthy controls were available in this cancer cohort). Each risk PRS was highly significantly associated with tumors of the respective cancer type (**Figure 5**), serving as a validation of both the scores and the imputed variants. Additionally, the smoking PRS was associated with lung tumors, and the pigment/sunburn PRS was associated with melanoma tumors as anticipated. No significant associations were observed for any mismatching PRS/cancer pairs, confirming empirically that our approach did not induce cancer-specific biases. This analysis demonstrates the potential power of tumor imputation to infer accurate PRSs and explore risk/exposure relationships within a clinical cohort.

**Figure 5:**
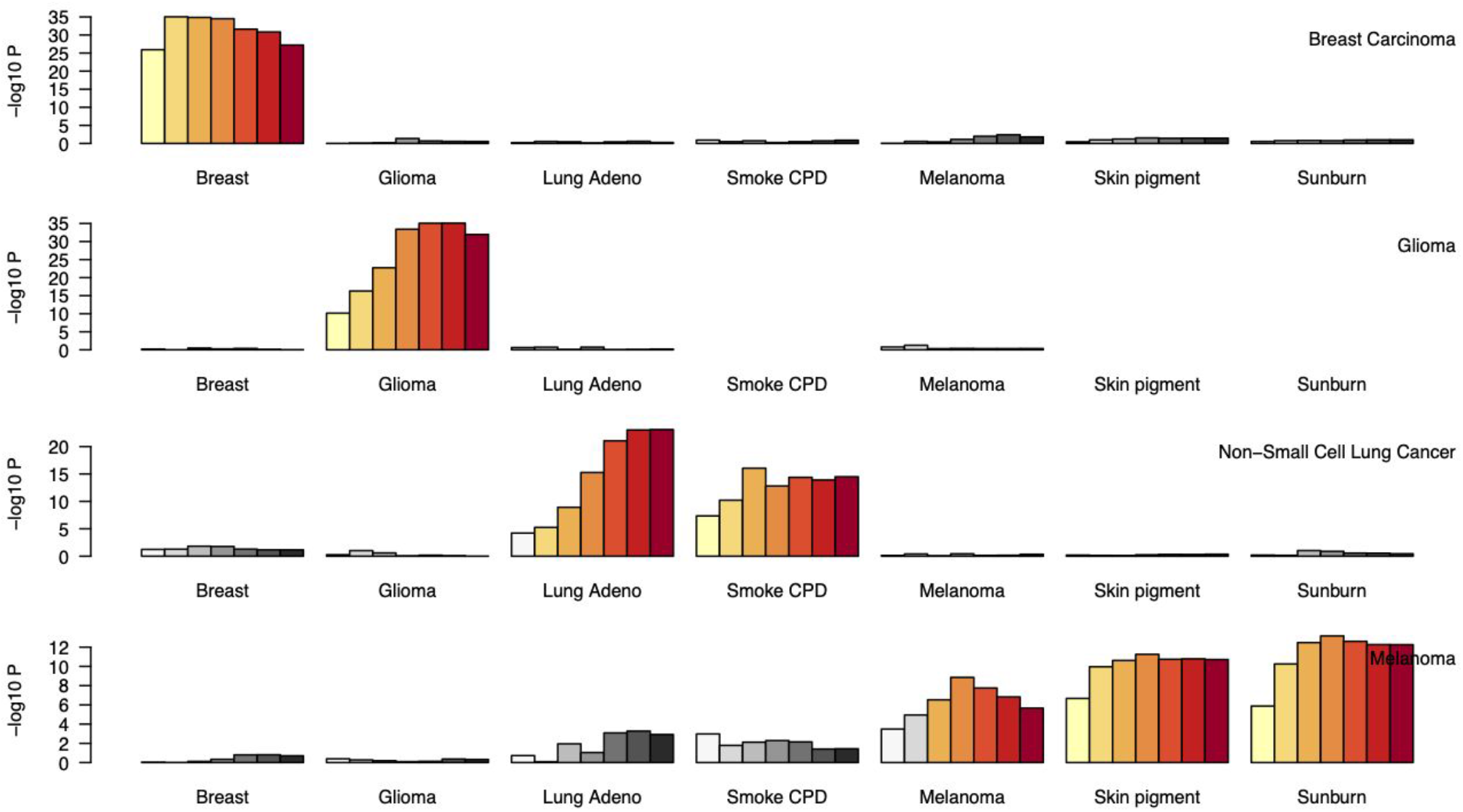
PRS associations with cancer types. Each column shows a different PRS type, subdivided by the PRS significance threshold (see Methods). Each row shows the true cancer type used as an outcome (cancer vs. rest) for association with PRS. Significance of the PRS-cancer type association is shown on the y-axis on the -log_10_ p-value scale (higher = more significant). Bonferroni significant associations are shown in orange shades, others are shown in gray. Note that significance of the PRS depends on both prediction accuracy as well as target sample size and should not be directly compared across cancer types.

### Inference of genetic ancestry

Germline variants can be used to discover and estimate genetic ancestry components, which are often partially distinct from self-reported race and ethnicity[33,34]. Previous work has shown that genetic data from an individual of unknown ancestry can be “projected” onto their ancestry component using weights from a reference panel [35] (akin to an ancestry PRS). We investigated this approach to ancestry inference by projecting tumor imputed genotypes into the principal components of three continental populations (European, African, and Asian; inferred from the 1000 Genomes Project reference data). The inferred ancestry revealed clines consistent with self-reported race (**Figure 6a**). Using the benchmarking samples, the correlation of ancestry estimates from tumor imputed variants versus germline genotyped variants was >0.98 with no significant outliers (**Figure 6b,c**). Continental ancestry was thus inferred from tumor imputed data with nearly perfect accuracy. We note that prior work showed ancestry projections from reference data are more accurate than in-sample principal component analysis[35], and thus we did not investigate the latter in the tumor data.

**Figure 6:**
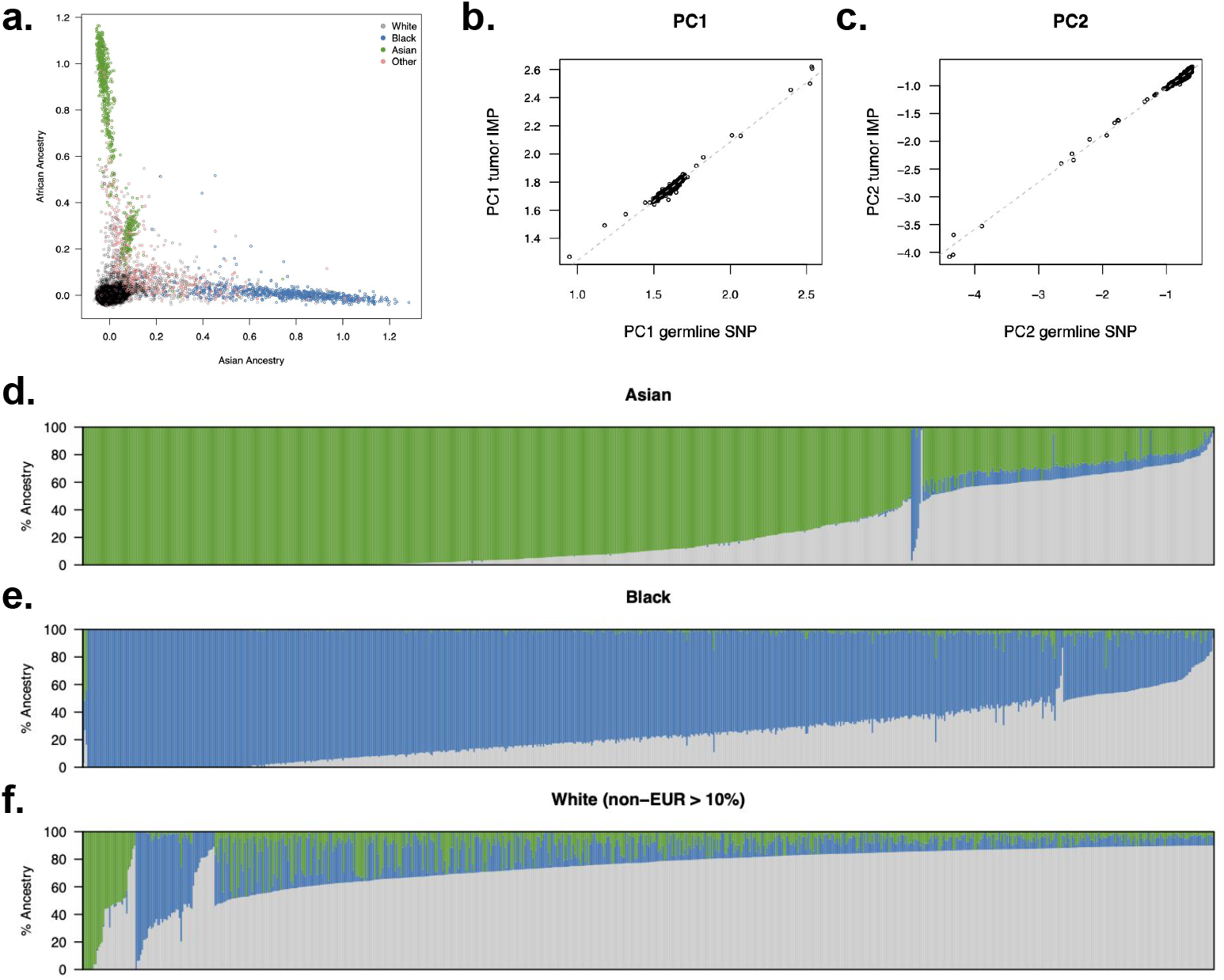
Genetic ancestry inferred from tumor sequencing. (**a**) Projected continental ancestry components for all individuals. Each dot is a projected sample (color-coded by self-reported race), x-axis is the Asian ancestry component and y-axis is the African ancestry component. (**b,c**) Correlation of imputed/genotyped ancestry component 1 and 2 respectively. (**d,e,f**) Ancestry fractions for individuals self-reporting as Asian, Black, and White (with >10% inferred non-European ancestry).

Having established accurate inference of genetic ancestry, we examined the variance in genetic ancestry across our full cohort of ∼25,000 samples. We first focused on individuals self-reporting as Asian (n=687) or Black (n=750) in the electronic health record (**Figure 6d,e**). As expected, self-reported Asian and Black patients exhibited predominantly East Asian and West African ancestry, respectively. However, admixture from other populations was also present in these samples: 50% of self-reported Asian patients had >10% ancestry from a non-Asian population; and 72% of self-reported Black patients had >10% ancestry from a non-African population. Next, we turned to self-reported White individuals with >10% non-European ancestry (n=730 out of 21,451 total self-reported Whites; **Figure 6f**). In addition to observing n=25 individuals with >50% East Asian ancestry and n=33 individuals with >50% African ancestry (possibly reflecting miscoded EHR data); we observed a cline of individuals with ancestry from both populations, likely encompassing Hispanic/Latino individuals who often exhibit this pattern of admixture[36] and self-report as White.

To demonstrate the relationship between genetic ancestry and somatic features, we focused on 2,900 non-small-cell lung cancer (NSCLC) patients with somatic, non-synonymous SNVs in the *EGFR* gene, which are known to have higher frequencies in Asians [37,38]. Restricting to individuals with Asian or European ancestry, we confirmed a highly significant increase in *EGFR* mutation rate in tumors from individuals with Asian ancestry (18% in samples of European ancestry; 57% in samples of Asian ancestry; p=3×10^−22^ by logistic regression). Genetic ancestry was more significantly associated with *EGFR* status than self-reported race (p=1.0×10^−3^ for ancestry and p=0.33 for race, in a joint model; **Table S2a**), highlighting the utility of the additional variation derived from inferred ancestry. Indeed, individuals that were self-reported White but had East Asian ancestry carried somatic *EGFR* SNVs at nearly the same rate as self-reported Asian individuals (47% and 58% respectively; **Table S2b**), serving as independent support of the inferred ancestry in these race/ancestry discordant samples.

## DISCUSSION

Traditionally, research cohorts for identifying germline and somatic cancer factors have been examined with different technologies and analysis methods, each optimized to specific goals. GWAS have identified over 1,000 SNPs associated with susceptibility to over 30 cancer types[39,40], but current GWAS techniques do not examine tumor sequences. Cancer-oriented cohorts like TCGA[41] obtained multiple functional data types, but sample sizes were too small for GWAS analyses, clinical data were sparse[42], and the patient populations had limited longitudinal follow-up. New multi-consortium studies promise to increase the sample size, but the high cost of employed technology (WGS) remains a limiting factor [15]. Targeted cancer sequencing thus offers a reduced cost, and the rich clinical data collected across hundreds of thousands of cancer patients[43] provide unprecedented opportunity for basic research and translational discovery. However, the lack of genome-wide germline data for these studies has limited the capacity to integrate GWAS-like analyses with somatic outcomes.

To overcome this gap between germline and somatic studies we implemented and validated an imputation framework to derive germline genotypes and downstream features directly from targeted tumor sequencing. Our pipeline offers the opportunity to run germline GWAS, estimate polygenic risk for complex phenotypes, and assign genetic ancestry. We demonstrated feasibility in real world data from >25,000 tumors, identifying highly-significant PRS associations and novel ancestry diversity. In particular, we found that ancestry scores were imputed with near perfect accuracy, providing a framework to easily incorporate genetic ancestry into the study of tumors from existing large-scale datasets and expand our knowledge of population-specific mechanisms[44]. Multiple emerging studies have demonstrated the utility of low-coverage sequencing of normal samples[17,21,22] and our work extends these findings to tumor-only sequencing data.

Our approach has several important limitations. First, while we show that ancestry and polygenic risk can be imputed with high and moderate-to-high accuracy (respectively) and negligible bias, individual variant calls should always be evaluated in the context of their imputation uncertainty. We observed a mean imputation correlation of 0.86 which, under standard assumptions of linearity, is expected to translate into an effective sample size of 0.86^2^ = 0.74x relative to direct germline genotyping[16]. While this is typically sufficient for GWAS discovery across thousands of samples, a wide range of accuracy was observed for individual variants, and associations with variants of high uncertainty must be interpreted with care. Second, while most genotyping errors will result in decreased sensitivity that may be acceptable in a discovery analysis, some forms of germline-somatic analysis may produce false positive associations. For example, we found that deep somatic deletions introduced noise and shifted the mean imputed variant towards the reference/common allele. This allele frequency shift may appear as a false positive association between the germline variant and a recurrent deletion (or other SCNAs correlated with the deletion). When testing individual variants in loss-prone regions, we therefore recommend either excluding individuals that carry a local deletion, or meta-analyzing carriers and non-carriers. Third, our study was limited to working with a single tumor-only panel sequencing platform (OncoPanel) which may not generalize to platforms used at other institutions. However, we did not observe substantial differences in performance across the three OncoPanel versions, which differed in their gene targets, and we anticipate that other panels targeting a similar range of genes (>200) with similar coverage distributions are likely to yield similar imputation accuracy; though broader evaluation is needed. For studies that perform tumor/normal matched sequencing, we expect that imputation from the normal sample would only further increase accuracy due to lack of confounding from somatic alterations (consistent with our observation that tumor purity slightly increased imputation error even in this tumor-only cohort). However, the optimal combination of tumor/normal data and sequencing platform for germline imputation remains an open question.

In addition to increasing sample sizes, the availability of germline genotypes in somatically phenotyped cohorts offers new avenues of research. Together with germline variation, rich clinical and omics measurements available across tens of thousands of samples will make it feasible to detect germline-somatic interactions, characterize the heritability of somatic events, tumor subtypes, disease prognosis, and therapy response, and identify germline biomarkers. To facilitate this research, code for all analyses described here is available in a repository and deployable workflow (see **Availability of data and materials**).

## CONCLUSIONS

In conclusion, we show using real data that common germline genotypes and derivative features can be accurately imputed from tumor-only panel sequencing. Our framework is publicly available and lays the path for germline studies from hundreds of thousands of tumors in existing datasets.

## Data Availability

The raw sequencing data are not publicly available because the research participant consent, privacy policy, and terms of service do not include authorization to share identifiable data.
The full analysis workflow is available at:
https://github.com/gusevlab/panel-imp
A containerized version of the imputation pipeline is available at: https://hub.docker.com/r/stefangroha/stitch_gcs

## FIGURE AND TABLE LEGENDS

**Figure S1**: (**a**) Histogram of broad cancer types in the full tumor cohort. (**b**) Histogram of coverage per sample across the full tumor cohort.

**Figure S2**: Histogram of imputation accuracy (Pearson correlation) using STITCH (gray) and GLIMPSE (red) imputation algorithms on chromosome 22.

**Figure S3**: Histogram of fraction of SNPs (y-axis) as a function of maximum absolute imputation error (x-axis) for all imputed SNPs (gray) and QC-passing SNPs (blue).

**Figure S4**: Histogram of imputation accuracy (Pearson correlation; shaded gray bars) and fraction of SNPs (black outlined bars) across QC thresholds. INFO: Imputation confidence score; HWE: Hardy-Weinberg equilibrium p-value; EAF: estimated allele frequency; PAF: estimated allele frequency based on reads.

**Figure S5**: Beehive plot of imputed HLA homozygosity probability (y-axis) versus true HLA homozygosity (x-axis) shown for MHC class I and class II. Homozygosity is defined as being homozygous for at least one allele.

**Figure S6**: Scatter plot of genotyped (x-axis) and imputed (y-axis) PRS scores across individuals (points). Scores used were: (a) breast cancer; (b) glioma; (c) smoking; (d) ovarian cancer; (e) prostate cancer; (f) kidney cancer; (g) lung cancer.

**Figure S7**: Violin plot of PRS error (genotyped minus imputed) across three versions of OncoPanel.

**Table S1**: Association of somatic features with imputation error. Estimates for a joint linear regression.

**Table S2**: (**a**) Association of *EGFR* somatic mutation carriers with race/ancestry features across three models. (**b**) *EGFR* somatic mutation carrier frequency across race/ancestry groups.

## DECLARATIONS

### Ethics approval and consent to participate

Written informed consent was obtained from participants prior to inclusion in this study (IRB approved protocols 11-104 and 17-000).

## Consent for publication

Not applicable.

## Availability of data and materials

The raw sequencing data are not publicly available because the research participant consent, privacy policy, and terms of service do not include authorization to share identifiable data.

The full analysis workflow is available at:https://github.com/gusevlab/panel-imp

A containerized version of the imputation pipeline is available at: https://hub.docker.com/r/stefangroha/stitch_gcs

## Authors’ contributions

AG and NZ conceived and designed the work; AG, SG, and KT contributed to the creation of new software in the work; all authors contributed to the acquisition, analysis, and interpretation of data as well as the drafting and revising of the work.

## Acknowledgements

The authors would like to acknowledge Bogdan Pasaniuc, Ethan Cerami, Guruprasad Ananda, Jian Carrot-Zhang, Laura MacConaill, Matthew Meyerson, Paz Polak, Siyang Liu, Sylvan Baca, and Yixuan He for helpful discussions and feedback. The authors would also like to acknowledge the DFCI Oncology Data Retrieval System (OncDRS) for the aggregation, management, and delivery of the clinical and operational research data used in this project; as well as the DFCI/BWH Data Sharing Group for the aggregation, management, and delivery of the genomics data used in this project.

