## Supplemental Material for "Constructing germline research cohorts from the discarded reads of clinical tumor sequences"

### Supplementary Figures

**Figure S1**

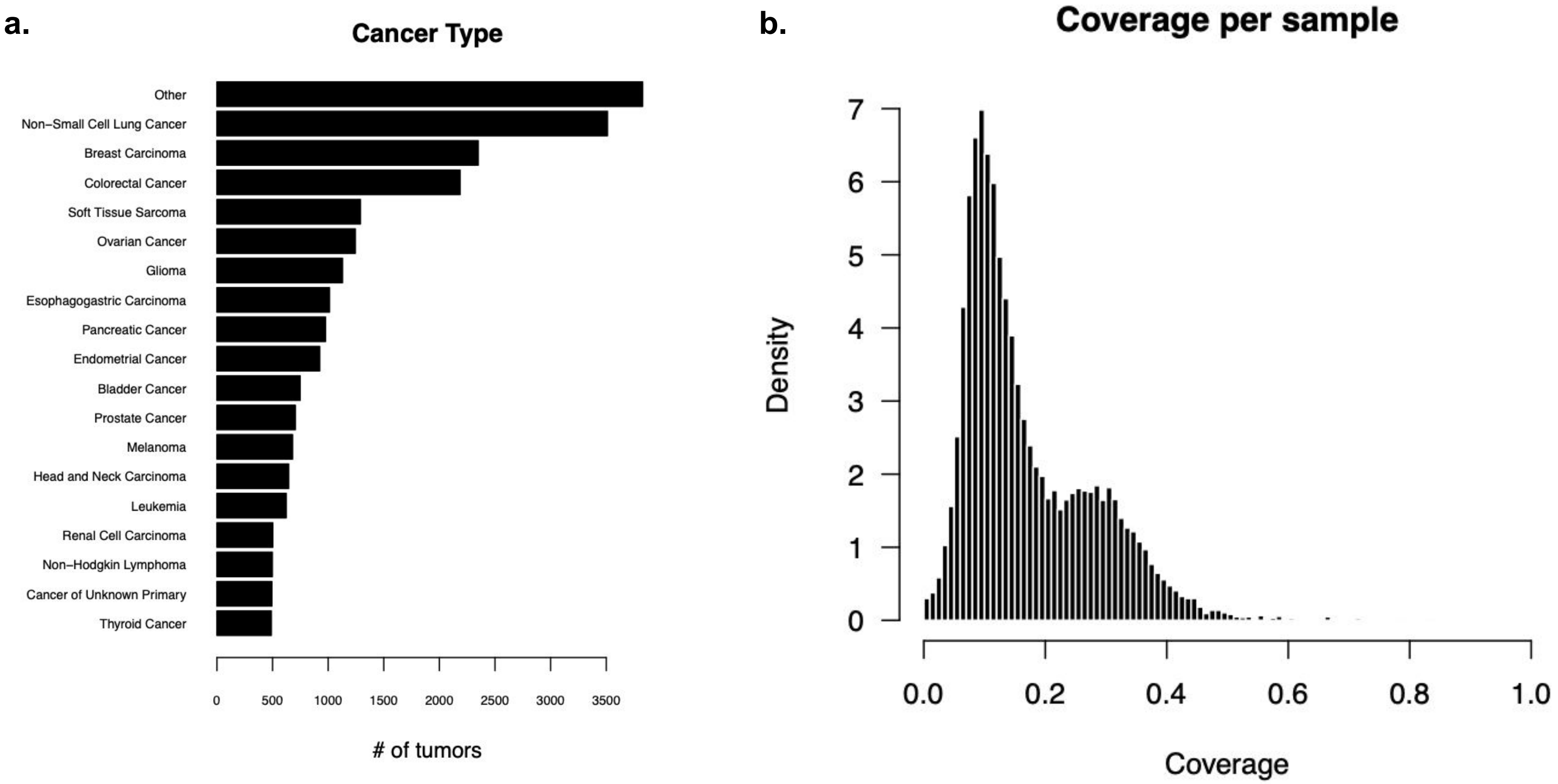
a.
Cancer Type
Other
Non−Small Cell Lung Cancer
Breast Carcinoma
Colorectal Cancer
Soft Tissue Sarcoma
Ovarian Cancer
Glioma
Esophagogastric Carcinoma
Pancreatic Cancer
Endometrial Cancer
Bladder Cancer
Prostate Cancer
Melanoma
Head and Neck Carcinoma
Leukemia
Renal Cell Carcinoma
Non−Hodgkin Lymphoma
Cancer of Unknown Primary
Thyroid Cancer
0
500
1000
1500
2000
2500
3000
3500
### of tumors
b.
Coverage per sample
Density
0
1
2
3
4
5
6
7
0.0
0.2
0.4
0.6
0.8
1.0
Coverage

**Figure S2**

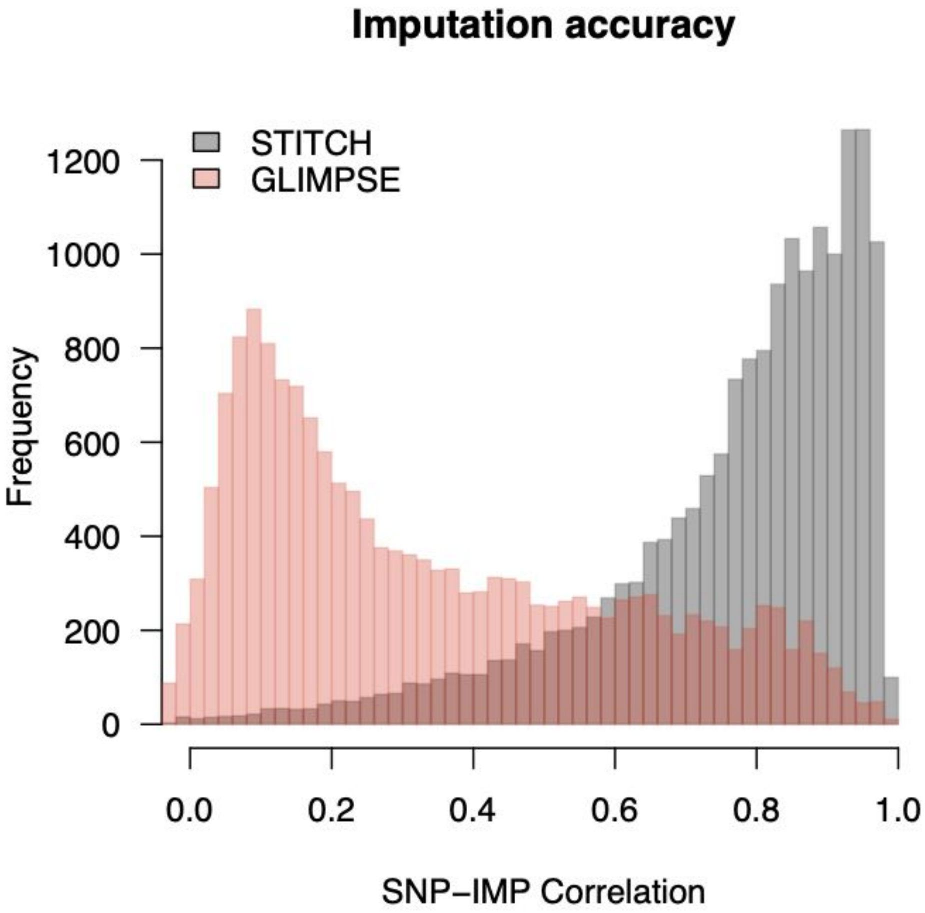

Imputation accuracy
STITCH
GLIMPSE
Frequency
0
200
400
600
800
1000
1200
0.0
0.2
0.4
0.6
0.8
1.0
SNP–IMP Correlation

**Figure S3**

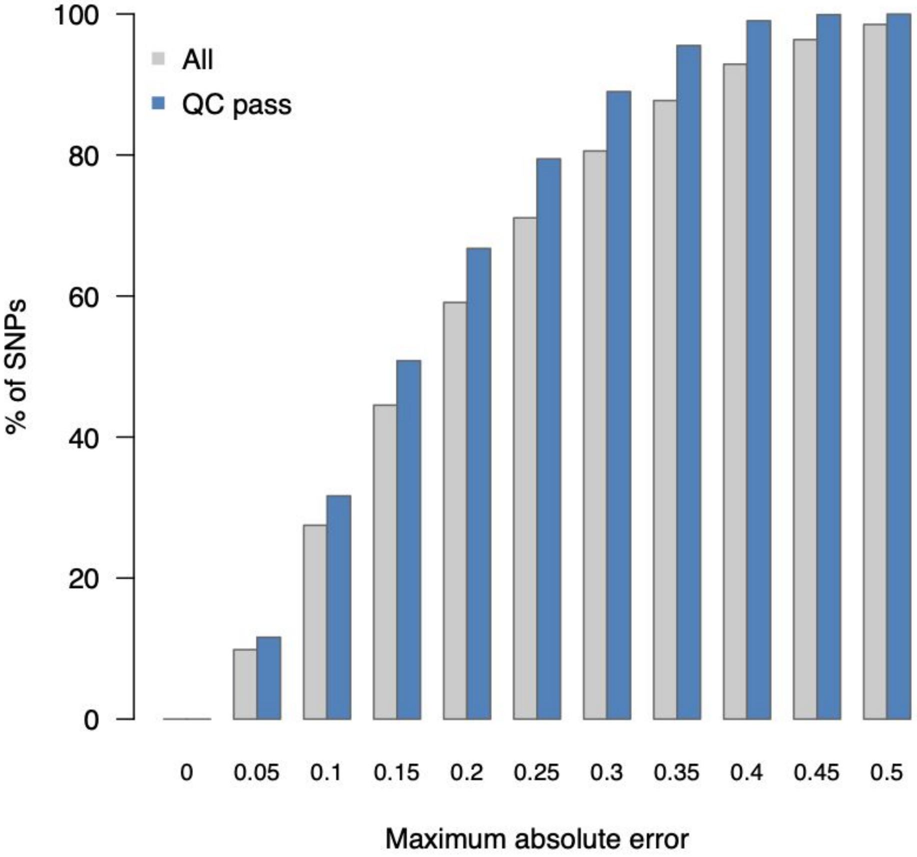

100
80
60
40
20
0
% of SNPs
All
QC pass
0
0.05
0.1
0.15
0.2
0.25
0.3
0.35
0.4
0.45
0.5
Maximum absolute error

### Figure S4

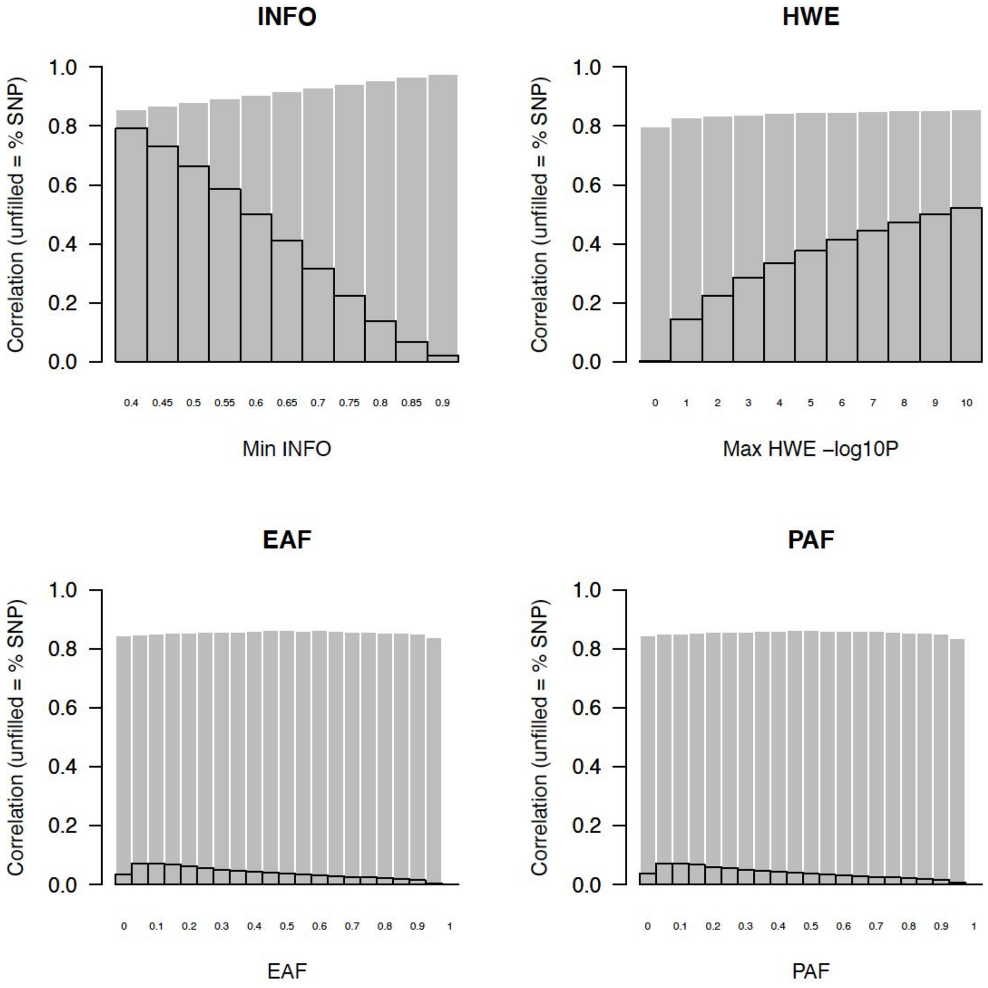
INFO
HWE
EAF
PAF
Correlation (unfilled = % SNP)
1.0
0.8
0.6
0.4
0.2
0.0
0.4 0.45 0.5 0.55 0.6 0.65 0.7 0.75 0.8 0.85 0.9
Min INFO
0 1 2 3 4 5 6 7 8 9 10
Max HWE –log10P
0 0.1 0.2 0.3 0.4 0.5 0.6 0.7 0.8 0.9 1
EAF
0 0.1 0.2 0.3 0.4 0.5 0.6 0.7 0.8 0.9 1
PAF

**Figure S5**

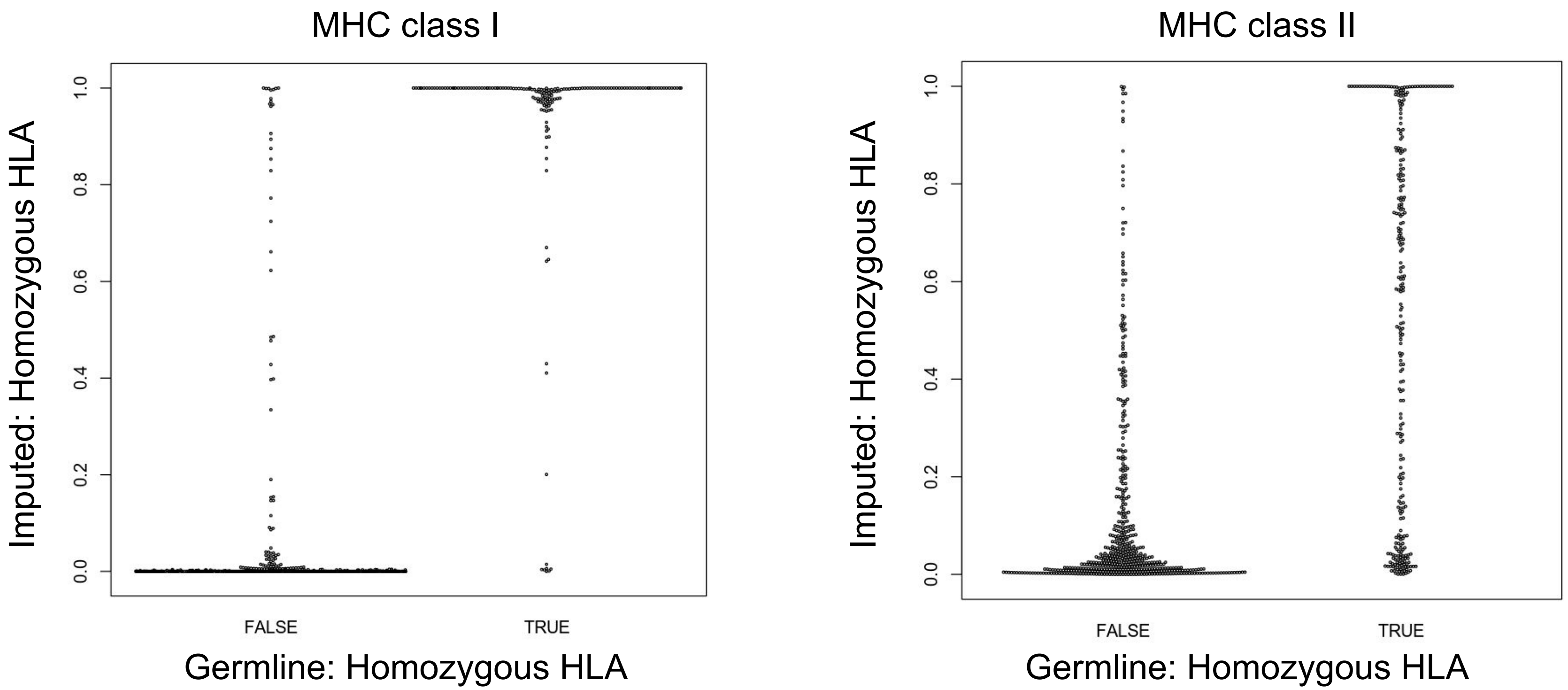
MHC class I
Imputed: Homozygous HLA
1.0
0.8
0.6
0.4
0.2
0.0
FALSE
TRUE
Germline: Homozygous HLA
MHC class II
Imputed: Homozygous HLA
1.0
0.8
0.6
0.4
0.2
0.0
FALSE
TRUE
Germline: Homozygous HLA

### Figure S6

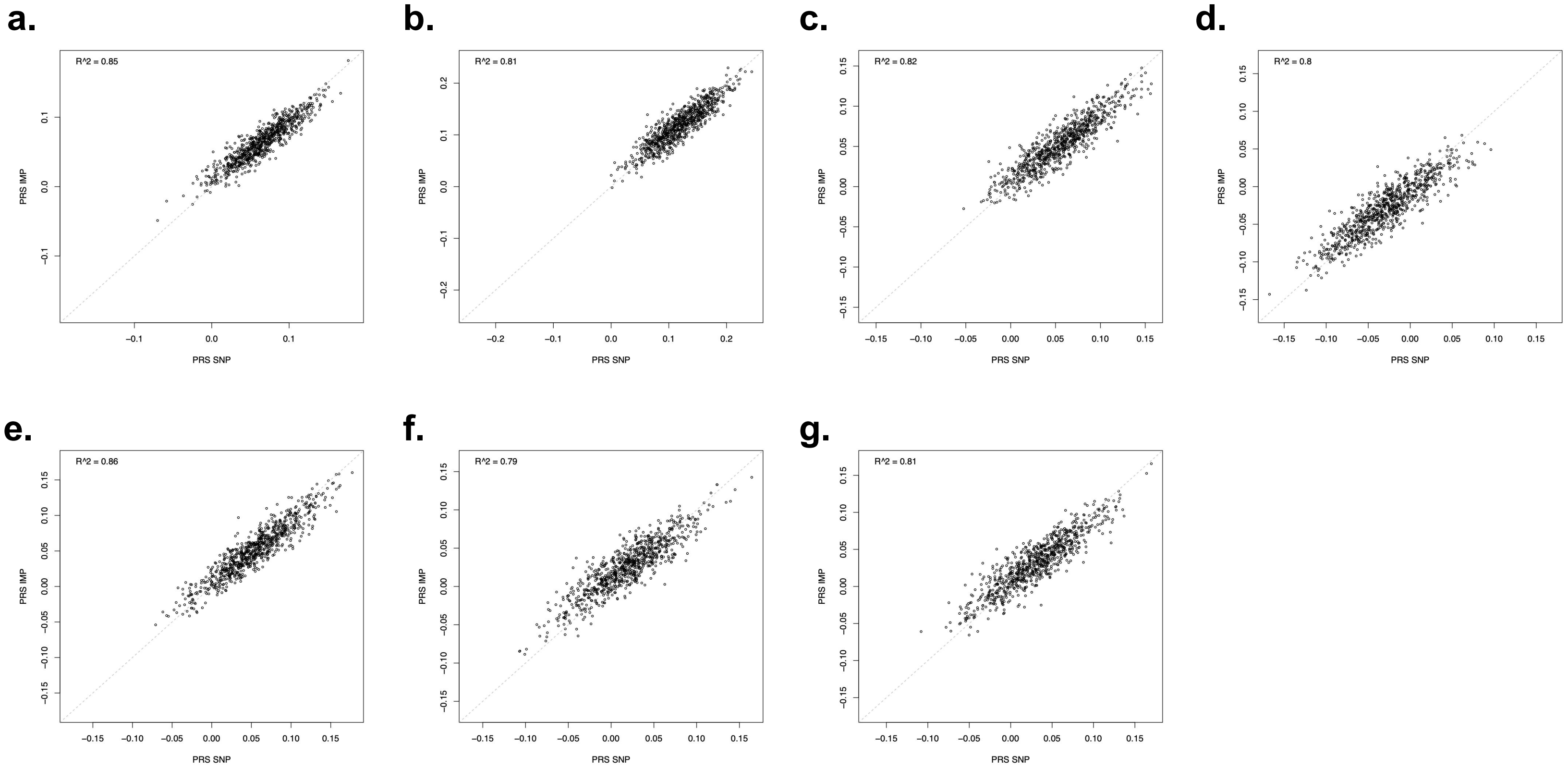
a.
R^2 = 0.85
PRS IMP
PRS SNP
b.
R^2 = 0.81
PRS IMP
PRS SNP
c.
R^2 = 0.82
PRS IMP
PRS SNP
d.
R^2 = 0.8
PRS IMP
PRS SNP
e.
R^2 = 0.86
PRS IMP
PRS SNP
f.
R^2 = 0.79
PRS IMP
PRS SNP
g.
R^2 = 0.81
PRS IMP
PRS SNP

### Figure S7

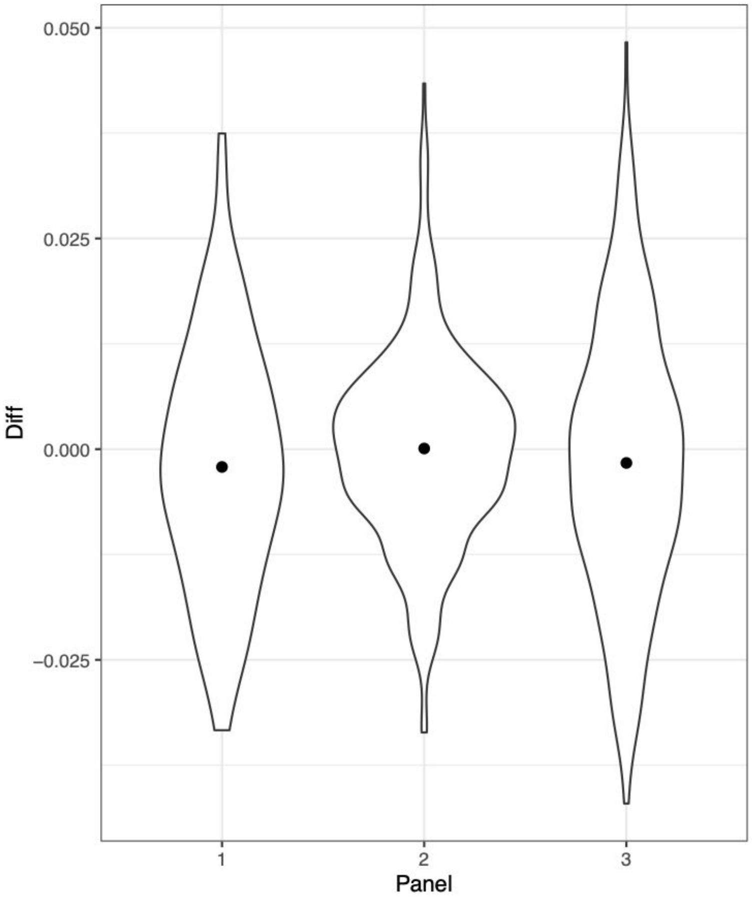

#### Table S1

| **Feature** | **Effect size** | **s.e.** | **T-statistic** | **P-value** |
|---|---|---|---|---|
| intercept | 1.76E-01 | 3.33E-03 | 5.29E+01 | 1.29E-264 |
| **Tumor purity >50%** | 5.51E-03 | 2.28E-03 | 2.41E+00 | 1.60E-02 |
| **Panel version == 2** | -2.85e-02 | 3.30E-03 | -8.66E+00 | 2.54E-17 |
| **Panel version == 3** | 2.90E-02 | 3.51E-03 | 8.28E+00 | 5.28E-16 |
| Metastatic tumor | 4.83E-03 | 2.73E-03 | 1.77E+00 | 7.68E-02 |
| Total Copy Number | 3.59E-03 | 9.63E-03 | 3.72E-01 | 7.10E-01 |
| Tumor Mutational Burden | 8.20E-05 | 1.45E-04 | 5.65E-01 | 5.72E-01 |

**Table S2a**

| Model | Ancestry p-value | Race p-value |
|---|---|---|
| EGFR ~ race | - | $2.3x10^{-21}$ |
| EGFR ~ ancestry | $3.5x10^{-22}$ | - |
| EGFR ~ ancestry + race | 0.001 | 0.33 |

**Table S2b**

| Race | All | Asian ancestry | European ancestry |
|---|---|---|---|
| Self-reported White | 17% | 47% | 17% |
| Self-reported Asian | 56% | 58% | - |
